# Mpox vaccination uptake in a UK community sample of gay, bisexual, and other men who have sex with men (GBMSM) the year following the 2022 Clade IIb mpox outbreak

**DOI:** 10.1101/2024.09.09.24313322

**Authors:** Dolores Mullen, Jessica Edney, Dawn Phillips, Ruth Wilkie, David Reid, Catherine M Lowndes, Erna Buitendam, Katy Sinka, Sema Mandal, Catherine H Mercer, John Saunders, Hamish Mohammed, Dana Ogaz

## Abstract

Mpox is an infectious disease transmitted through close contact. It is caused by the monkeypox virus, which is endemic to some countries of West and Central Africa. A multi-country outbreak of mpox occurred in 2022, and the UK experienced rapid community transmission associated with sexual contact, mainly, but not exclusively among networks of gay, bisexual, and other men who have sex with men (GBMSM). In response to the outbreak in the UK, a reactive mpox vaccination programme was targeted to those most at risk. We explore the uptake and course completion of mpox vaccination in GBMSM taking part in an online survey in 2023. Findings from this community sample indicate vaccination uptake in around two-thirds of participants meeting mpox proxy eligibility criteria with high levels of course completion among all and eligible participants that were ever vaccinated. Vaccine non-offer was a barrier to uptake, as nearly a third of those eligible but unvaccinated reported never having received an mpox vaccine offer. Continued targeting of vaccination to GBMSM at highest risk of mpox at SHS, with community-support, will help facilitate equitable uptake of vaccination.

## Scientific Letter

Control of the 2022 Clade IIb mpox outbreak in the UK, primarily affecting gay, bisexual, and other men who have sex with men (GBMSM), was achieved through behavioural change and targeted vaccination, both supported by coproduced health promotion (1, 2). In November 2022, the Joint Committee on Vaccination and Immunisation recommended a routine mpox vaccination programme targeted primarily to GBMSM (3). We report mpox vaccination uptake and course completion among a community sample of GBMSM a year following the 2022 mpox outbreak to complement prior findings on mpox vaccination uptake (1).

Using data collected from the ‘Reducing inequalities in Sexual Health’ (RiiSH) 2023 survey (November-December 2023), part of a series of cross-sectional surveys examining the sexual health and wellbeing of a GBMSM community sample (4), we performed descriptive analyses of vaccination offer and uptake (ever having ≥1, 2 doses) (%, 95% CI), vaccination course completion (2 doses), recency of last dose (June 2022 to survey completion), and method of administration (subcutaneous, intradermal, both) among all participants and those eligible for vaccination. Vaccine eligibility was based on national guidance (5) and defined in the analysis using the following proxy criteria (any of the following since August 2023): ≥10 physical male sex partners, meeting a male sex partner(s) in a sex-on-premises venue, sex party, or cruising ground; a positive STI test; and/or in the last year, report of: HIV pre-exposure prophylaxis use; or use of recreational drugs associated with chemsex (crystal methamphetamine, mephedrone or gamma-hydroxybutyrate/gamma-butyrolactone).

Of 1,106 participants (median age 44 years [IQR:34-54]; 89% White ethnicity; 78% UK-born; 78% employed; 62% degree-level education; 24% reporting ≥10 physical male sex partners in the last 3 months), 58% (636/1,106) were eligible for mpox vaccination.

Among eligible participants, 66% (419/636) were ever offered an mpox vaccine. Uptake of ≥1 mpox vaccine doses was 94% (393/419) among those offered, or 62% (95% CI: 56%-68%, 393/636) in all eligible participants. A minority of eligible participants reported vaccine offer but declined uptake (6%, 26/419). Nearly one-third of eligible participants reported never having received an mpox vaccination offer (29%, 185/636) of which, 65% (121/185) had visited a sexual health service (SHS) in the last year. In those eligible and vaccinated with ≥1 dose, 77% (95% CI: 69%-87%, 304/393) reported course completion.

In all participants, 46% (508/1,106) were offered an mpox vaccine. Uptake of ≥1 mpox vaccine dose was 93% (470/508) among those offered, or 42% (95% CI: 39%-47%, 470/1,106) in all participants. Most participants reporting ≥1 mpox vaccine doses met proxy eligibility criteria (84%, 393/470). In participants vaccinated with ≥1 dose, 76% (95% CI: 68%-84%, 356/470) reported course completion. Among those with an incomplete vaccination (n=114, only 1 dose), 90% (103/114) were vaccinated before August 2023. In all those vaccinated, most reported exclusive subcutaneous vaccination (68% [77/114] in those with one dose; 50% [179/356] in those with 2 doses).

Findings from this community sample indicate vaccination uptake in around two-thirds of participants meeting mpox proxy eligibility criteria with high levels of course completion among all and eligible participants that were ever vaccinated. Vaccine non-offer was a significant barrier to uptake, as nearly a third of those eligible but unvaccinated reported never having received an mpox vaccine offer. Most participants (over 90%) who were ever offered an mpox vaccine reported uptake, but the reasons for not accepting the offer of vaccination were not collected in the survey and may be due to multiple factors such individuals’ assessment of their level of risk (6). Given the cross-sectional design of the RiiSH survey, we cannot determine the temporality of vaccination offer in relation to most sexual risk behaviours comprising our vaccination eligibility proxy; however, we found that most participants who did report vaccination met proxy criteria. While RiiSH participants may represent a sample with higher health literacy and social mobility relative to nationally representative GBMSM survey samples (7), our findings represent key populations that are targeted for mpox vaccination where, like similar convenience samples, participants are more likely to report sexual risk behaviours than the general GBMSM population (8).

Lessons learned from the 2022 mpox outbreak highlight the importance of a community-supported response and rapid deployment of interventions as part of a series of combination prevention tools. Further studies are needed to understand the individual- and structural-level barriers to mpox vaccination initiation and completion. Continued targeting of vaccination to GBMSM at highest risk of mpox at SHS, with community-support, will help facilitate equitable uptake of vaccination.

## Supporting information

Mpox vaccination and offer reported in a) all and b) vaccine eligible RiiSH 2023 participants, November/December 2023

## Data Availability

Data availability statement:
The data that support the findings of this study are available upon reasonable request from the UK Health Security Agency (UKHSA). Requests can be directed to.

## Acknowledgements

The authors thank all the participants involved in this study.

