## Supplementary material for "Mpox vaccination uptake in a UK community sample of gay, bisexual, and other men who have sex with men (GBMSM) the year following the 2022 Clade IIb mpox outbreak": Mpox vaccination and offer reported in a) all and b) vaccine eligible RiiSH 2023 participants, November/December 2023

a) All participants

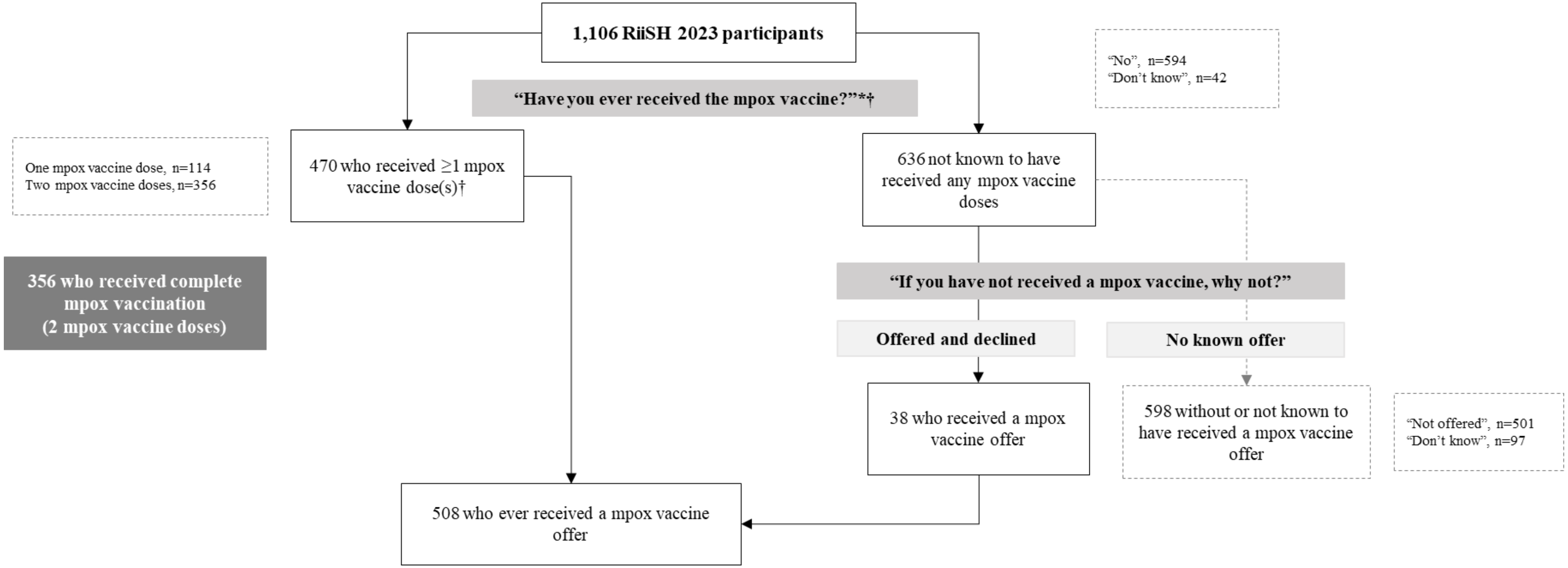


*Question preamble: "Mpox (formerly known as 'monkeypox') is a disease caused by infection with the monkeypox virus. Mpox vaccination usually involves having two doses, given by injection, at the recommended intervals." †Vaccine offer assumed in those reporting ≥1 vaccine dose(s).

b) Vaccine eligible participants

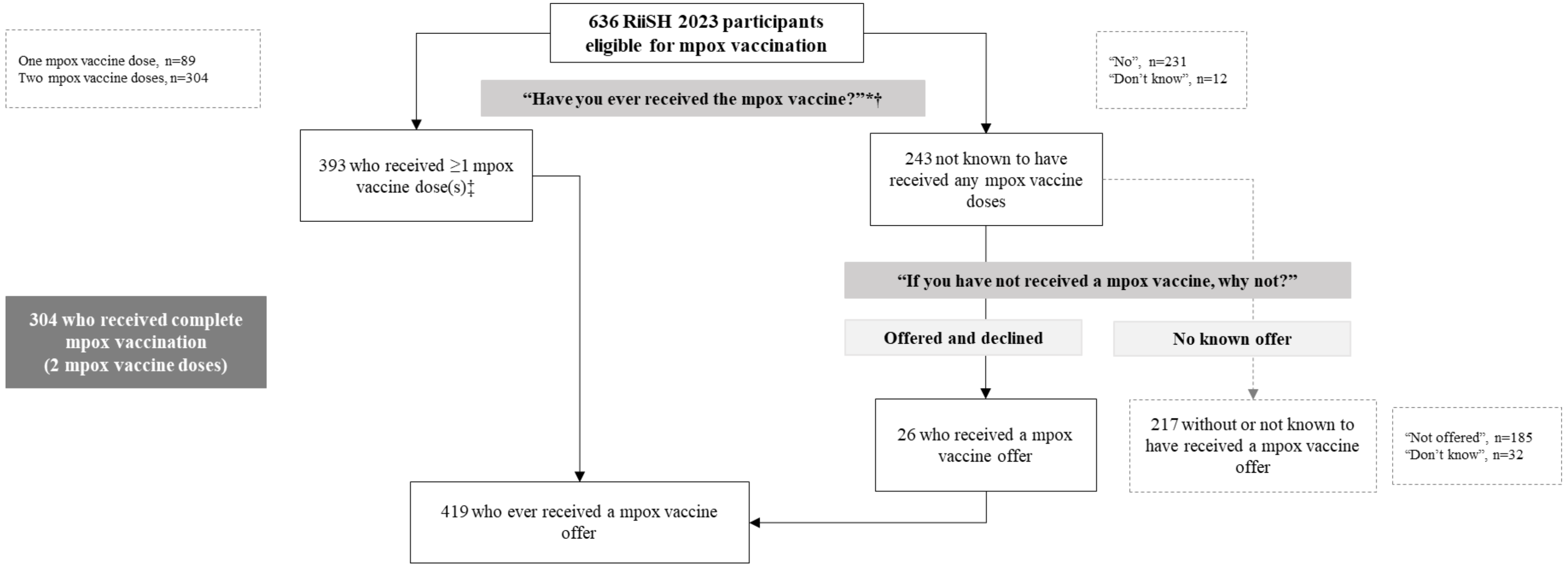


Vaccine eligibility was defined using proxy measures in line with national guidance comprising any of the following since August 2023: ≥10 physical male sex partners, meeting a male sex partner(s) in a sex on premises venue, sex party, or cruising grounds; a positive STI test; and/or in the last year, report of: HIV pre-exposure prophylaxis use; or use of recreational drugs associated with chemsex (crystal methamphetamine, mephedrone or gamma-hydroxybutyrate/gamma-butyrolactone); excludes those that were vaccinated but not considered eligible (n=77, 16.4% of all vaccinated [77/470]). *Question preamble: "Mpox (formerly known as 'monkeypox') is a disease caused by infection with the monkeypox virus. Mpox vaccination usually involves having two doses, given by injection, at the recommended intervals." † Vaccine offer assumed in those reporting ≥1 vaccine dose(s).
